# Predicting Nicotine Metabolism Across Ancestries Using Genotypes

**DOI:** 10.1101/2021.12.19.21267947

**Authors:** James W Baurley, Andrew W Bergen, Carolyn M Ervin, Sung-shim Lani Park, Sharon E Murphy, Christopher S McMahan

**Affiliations:** BioRealm LLC, 340 S Lemon Ave, Suite 1931, 91789 Walnut, CA, USA; Clemson University, 220 Parkway Drive, 29634 Clemson, SC, USA; Oregon Research Institute, 1776 Millrace Drive, 97403 Eugene, OR, USA; University of Hawaii, 701 Ilalo Street, 96813 Honolulu, HI, USA; University of Minnesota, 2231 6th St SE, 55455 Minneapolis, MN, USA

**Keywords:** nicotine metabolism, smoking cessation, machine learning, statistical learning, polygenic risk score, nicotine biomarkers

## Abstract

**Background:** There is a need to match characteristics of tobacco users with cessation treatments and risks of tobacco attributable diseases such as lung cancer. The rate in which the body metabolizes nicotine has proven an important predictor of these outcomes. Nicotine metabolism is primarily catalyzed by the enzyme cytochrone P450 (CYP2A6) and CYP2A6 activity can be measured as the ratio of two nicotine metabolites: *trans*-3’-hydroxycotinine to cotinine (NMR). Measurements of these metabolites are only possible in current tobacco users and vary by biofluid source, timing of collection, and protocols; unfortunately, this has limited their use in clinical practice. The NMR depends highly on genetic variation near *CYP2A6* on chromosome 19 as well as ancestry, environmental, and other genetic factors. Thus, we aimed to develop prediction models of nicotine metabolism using easy to obtain genotypes and individual characteristics.

**Results:** We identified four multiethnic studies with nicotine metabolites and DNA samples. We constructed a 263 marker panel from filtering genome-wide association scans of the NMR in each study. We then applied seven machine learning techniques to train models of nicotine metabolism on the largest and most ancestrally diverse dataset (N=2239). The models were then validated out-of-sample in the other three studies (total N=1415). Using cross-validation, we found the correlations between the observed and predicted NMR ranged from 0.69 to 0.97 depending on the model. When predictions were averaged in an ensemble model, the correlation was 0.81. The ensemble model generalizes well out-of-sample across ancestries, despite differences in the measurements of NMR between studies, with correlations of: 0.52 for African ancestry, 0.61 for Asian ancestry, and 0.46 for European ancestry. The most influential predictors of NMR identified in more than two models were rs56113850, rs11878604, and 21 other genetic variants near *CYP2A6* as well as age and ancestry.

**Conclusions:** We have developed an ensemble of seven models for predicting the NMR across ancestries from genotypes and age, gender and BMI. Predictions from these models validate out-of-sample in three datasets and associate with nicotine dosages. The knowledge of how an individual metabolizes nicotine could be used to help select the optimal path to reducing or quitting tobacco use, as well as, evaluating risks of tobacco use.

## 1 Background

Tobacco smoking is a leading cause of global preventable disease and death. Nicotine, the component of tobacco that sustains nicotine addiction, makes tobacco smoking highly addictive and difficult to quit. Nicotine is primarily metabolized by the cytochrome P450 2A6 (CYP2A6) enzyme. Individual variations in CYP2A6 activity have been found to influence smoking behaviors [1]. A biomarker for measuring CYP2A6 enzymatic activity is the nicotine metabolite ratio (NMR), the ratio of two nicotine metabolites, *trans*-3’-hydroxycotinine (3HC) to cotinine (COT) [2]. The NMR has been shown to be associated with smoking behaviors [1], smoking dose and risk of lung cancer[3], alcohol consumption [4], and smoking cessation [5]. As a result, there have been repeated calls for screening based on the NMR [6].

There remain technical challenges in measuring nicotine metabolism that limits its potential clinical use. Ideally measurement would involve controlled nicotine administration trials which is simply not feasible in large population based studies or in screening. Biochemical measurement of the NMR requires serum, plasma, saliva, or urine for analyte analysis. While measurements of nicotine metabolites have good reproducibility within these biofluids [7], differences in metabolite measurements (e.g. total or free 3HC and COT) make results difficult to compare and interpret across studies [8]. Biochemical measurement of the NMR also requires biofluids to be collected relatively soon after the intake of nicotine, which is impractical in former smokers or if a smoker is an occasional user of tobacco. This limits risk assessments of tobacco attributable diseases and comorbidities to current smokers.

Genomic prediction of the NMR is a promising alternative to direct biochemical measurements. Genotyping services are widespread and feasible within clinical laboratories. A number of functional variants of *CYP2A6* have been shown to be associated with the NMR [9]. More recently genome-wide genotyping have identified additional variants that are associated with the NMR [10, 11, 12, 13]. Genomic data has been used to build more comprehensive genomic models [14] and polygenic risk scores [15] for predicting nicotine metabolism. But these models do not generalize across ancestries, requiring the development of ancestry-specific or transferable risk scores [16].

In this work we present the development and validation of an ensemble of models trained to predict NMR using genotypes and basic covariates across ancestries. We begin by prioritizing genetic markers found to be associated with the NMR in four multiethnic studies. We then apply an ensemble of machine learning algorithms to the largest study to train models which are then assessed directly in the other three by comparing observed to predicted NMR. The resulting selected variables and validated models can be used to assess nicotine metabolism in current or former tobacco users. This knowledge can help inform clinical decisions making on the optimal path to smoking cessation and communicate risks for tobacco-related outcomes.

## 2 Methods

### 2.1 Source of data

Four studies were identified with measured NMR or metabolite data, genomic data or DNA samples available for genotyping, and basic demographic variables. These four studies were used for training and validation of NMR models. They are summarized below.

#### 2.1.1 Multiethnic Cohort, MEC

The MEC was established in Hawaii and California (primarily Los Angeles County) to study diet and cancer in the United States [17, 18]. From 1993 to 1996, individuals of both sexes, aged 45-75, and from five major racial/ethnic groups (Latino, African-American, Japanese-American, White, Native Hawaiian) were recruited. Participants completed a baseline questionnaire of demographic characteristics, anthropometrics, smoking history and other lifestyle factors. This study uses a subcohort of 2,239 lung cancer free participants who were current smokers at time of biospecimen collection [19]. Collected biospecimens include blood and urine (overnight for Hawaii or first morning in California). Nicotine metabolites were quantified using liquid chromatography–tandem mass spectrometry [20]. Nicotine equivalents (NE) was defined as the sum of total nicotine, total COT, and total 3HC. Here total refers to the sum of the compound and its glucuronide conjugate. The NMR was defined as the urinary total 3HC to free COT ratio.

#### 2.1.2 Center for the Evaluation of Nicotine in Cigarettes, CENIC

CENIC conducted studies of the effects of reduced nicotine cigarettes on smoking outcomes. 550 participants across 8 United States institutions were randomized to one of seven nicotine levels between June 2013 and July 2014 [21] and had DNA available for analysis. Participants were adult daily smokers that smoke an average of at least five cigarettes per day for at least one year and had either a cotinine level > 100 ng/mL or expired carbon monoxide of > 8 ppm [21]. Smokers were initially assessed using their usual brand cigarettes. Nicotine metabolites were measured with liquid chromatography with tandem mass spectrometry. Total nicotine equivalents (TNE) was defined as the sum of total nicotine, total COT, total 3HC, and N-oxide. The NMR was defined as the urinary total 3HC to free COT ratio.

#### 2.1.3 Hawaii Smokers Study, HSS

The HSS was comprised of 600 participants randomly selected from the MEC participants who were current smokers at time of study, reporting that they smoked at least 10 cigarettes per day, had no history of cancer, and were self-reported Japanese, European, or Hawaiian ancestry [22]. Study interview and blood and 12-hour urine samples were collected independent of the previously mentioned MEC biospecimen collection. Analysis of total urinary nicotine, COT, and 3HC concentration was done by GC/MS (gas chromatography/mass spectrometry). NE was defined as the sum of nicotine, COT, and 3HC. The NMR was defined as the urinary total 3HC to total COT ratio.

#### 2.1.4 Laboratory studies of nicotine metabolism, METS

The METS included 315 unrelated African-American, Asian-American, and European-American individuals from three laboratory studies of nicotine metabolism [12]. The studies included Pharmacokinetics in Twins (PKTWIN) [23], Pharmacogenetic Study of Nicotine Metabolism (588) [24], and SMOking in FAMilies (SMO-FAM) [25]. Blood or saliva was collected 6 hours after the administration of labeled nicotine and cotinine in smokers and non-smokers. Nicotine metabolite levels were assessed using gas chromatography-tandem mass spectrometry methods. The NMR was defined as the 6 hour plasma or saliva 3HC to COT ratio.

### 2.2 Response Variable

We aim to develop a predictive model of the urinary nicotine metabolite ratio (the ratio of total 3HC to free COT) from genotypes and covariates.

### 2.3 Predictors

#### 2.3.1 Genotypes

Imputed genotypes were already available for the MEC and METS studies [13, 12]. The MEC was genotyped on the Illumina Human1M-Duo BeadChip. The METS were previously genotyped on the Smokescreen Genotyping Array [26]. Both were imputed to include variants in the 1000 Genomes Project reference populations using standard phasing and imputation best practices at the time [13, 12].

DNA samples from HSS and CENIC were genotyped specifically for this project on the Smokescreen Genotyping Array. 200 ng of genomic DNA were plated using Axiom 2.0 Reagent Kits and processed on the GeneTitan MC instrument. Analysis of the raw data was performed using Affymetrix Power tools (APT) v-1.16. Additional steps were performed using SNPolisher to identify and select probe sets and high quality variants for downstream analysis. Quality control steps for samples included comparisons of self-reported and genomic gender and ancestry, detection of excessive heterozygosity (> 0.20), genotype concordance among known duplicates, and removal of unexpected duplicates and related samples. Quality control steps for genetic variants included missingness > 5% and deviation from Hardy Weinberg equilibrium (*p <*1E-10). After quality control, HSS had genotypes for 585 individuals on 569,986 genetic variants. CENIC had genotypes for 515 individuals on 570,258 genetic variants.

We used genome-wide imputation to harmonize genotypes across the studies prior to analysis. Alleles were reported on the forward strand and conform-gt was used to ensure consistency with the 1000 Genomes Phase 2 version 5a data files prepared for use with the Beagle imputation software. Beagle 5.2 was used to phase genotypes and impute ungenotyped or missing genotypes [27]. The resulting genotype dosages for variants typed on the Smokescreen Genotyping Array were imported into a Postgres database.

#### 2.3.2 Covariates

We compiled age, sex, self-identified ethnicity, body mass index (BMI), and smoking status (from METS) from study datasets. Additionally, ancestry proportions were estimated by extracting genotypes for 5516 ancestry informative markers from the study data and combining it with genotypes from 1000 Genomes Project Phase 3 version 5a. fastSTRUCTURE was used with default settings and *k* = 3 populations [28]. Populations assignments from the 1000 Genome Project and self-reported race from the studies were used to label the estimated European, Asian, and African ancestry proportions.

### 2.4 Sample size

The NMR was merged with genotypes and covariates for each study to create the analytic dataset. Sample sizes were 2,239 for MEC, 515 for CENIC, 585 for HSS, and 315 for METS.

### 2.5 Missing data

HSS was missing 5 observations for NMRs and those records were dropped from the analysis. The genome-wide assocation scans of NMR used for marker nomination used complete observations. In model training and validation, missing values were imputed using the missMDA package in R [29]. Briefly, the data for candidate predictors were stacked across studies, the number of dimensions were estimated by principal components analysis (PCA), and the missing values were imputed with the PCA model.

### 2.6 Statistical analysis methods

#### 2.6.1 Marker Nomination

Prior to training NMR prediction models, we nominated markers to consider using results from genome-wide association scans. The scans used models of the form

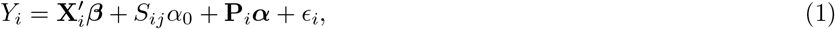

where *Y*_*i*_ is the natural log NMR measured on the *i*th individual, **X**_*i*_ is a vector of covariates with ***β*** being the corresponding vector of regression coefficients, *S*_*ij*_ is the genetic variant under consideration with *α*_0_ being the corresponding regression coefficient, **P**_*i*_ is a vector of principal components computed on the genotypes design matrix with ***α*** being the corresponding vector of regression coefficients, and *ϵ*_*i*_ is the error term. In this analysis, we controlled for age, sex, ancestry, body mass index (BMI), and smoking status in the METS. We used the first 50 principal components of the genotype design matrix to account for genetic relatedness/ancestry among the study participants. The p-values for the test of *H*_0_ : *α*_0_ = 0 vs. *H*_1_ : *α*_0_ ≠ 0 were computed.

From these results, we selected 200 genetic variants from each study based on the smallest p-values with allele frequencies > 1%. We took the union of these sets and retained genetic variants with evidence (*p <* 0.05) of association with the NMR in at least two of the four studies.

#### 2.6.2 Model Training and Validation

To develop a predictive model of NMR, we took an ensemble based approach that leveraged a suite of machine learning algorithms. This suite consisted of partial least squares [30], projection pursuit [31], elastic net [32], support vector machine (with a linear and radial basis function kernel) [33], gradient boosting machine [34], and random forests [35]. Each of these machine learning models was fit to the MEC data (the largest and most diverse of the four studies), treating the *Y*_*i*_ (NMR measured on the *i*th subject) as the response variable. To explain the heterogeneity in the NMR in this analysis, we used a feature set consisting of age, gender, BMI, Asian and African ancestry proportions, and the 263 prioritized markers arising from the marker nomination step. For notational brevity, we denote the feature set for the *i*th observation by **F**_*i*_. The R package caret was used to fit and train all of the models using the methods listed in Table 5.3.

It is important to note that the fitting process for each of the aforementioned models required the selection of tuning parameters that have to be specified in a methodical way to avoid issues of over- and under-fitting the data. Classically, this issue can also be described via the bias-variance tradeoff. That is, a model that is not appropriately regularized, or is overspecified, can over-fit the data thus reducing the bias at the expense of increased variability. In contrast, an underspecified model, or over regularized, could provide for less variable predictions but at the expense of increased bias. To choose the tuning parameters, we implemented repeated (10 times) 10-fold cross validation at an array of candidate tuning parameter values; for more on repeated cross validation see [36]. The grid of tuning parameters were designed based on initial analyses based on default settings, prior experience, and to ensure that the optimal configuration existed on the interior of the grid; i.e, the optimal value did not exist on the boundary of the grid. The optimal tuning parameter configuration for each of the machine learning models was determined to be the one that minimized the cross-validation error. Table 5.3 summarizes the candidate tuning parameters for each of the machine learning models. Note, our cross validation strategy requires fitting 100 models for each tuning parameter configuration for each of the considered machine learning model. Figure 1 provides a summary of these fits at their optimal tuning parameter configuration. This summary includes the mean absolute error (MAE), the root mean squared error (RMSE), and the R-squared value for each of the 100 fits.

**Figure 1.**
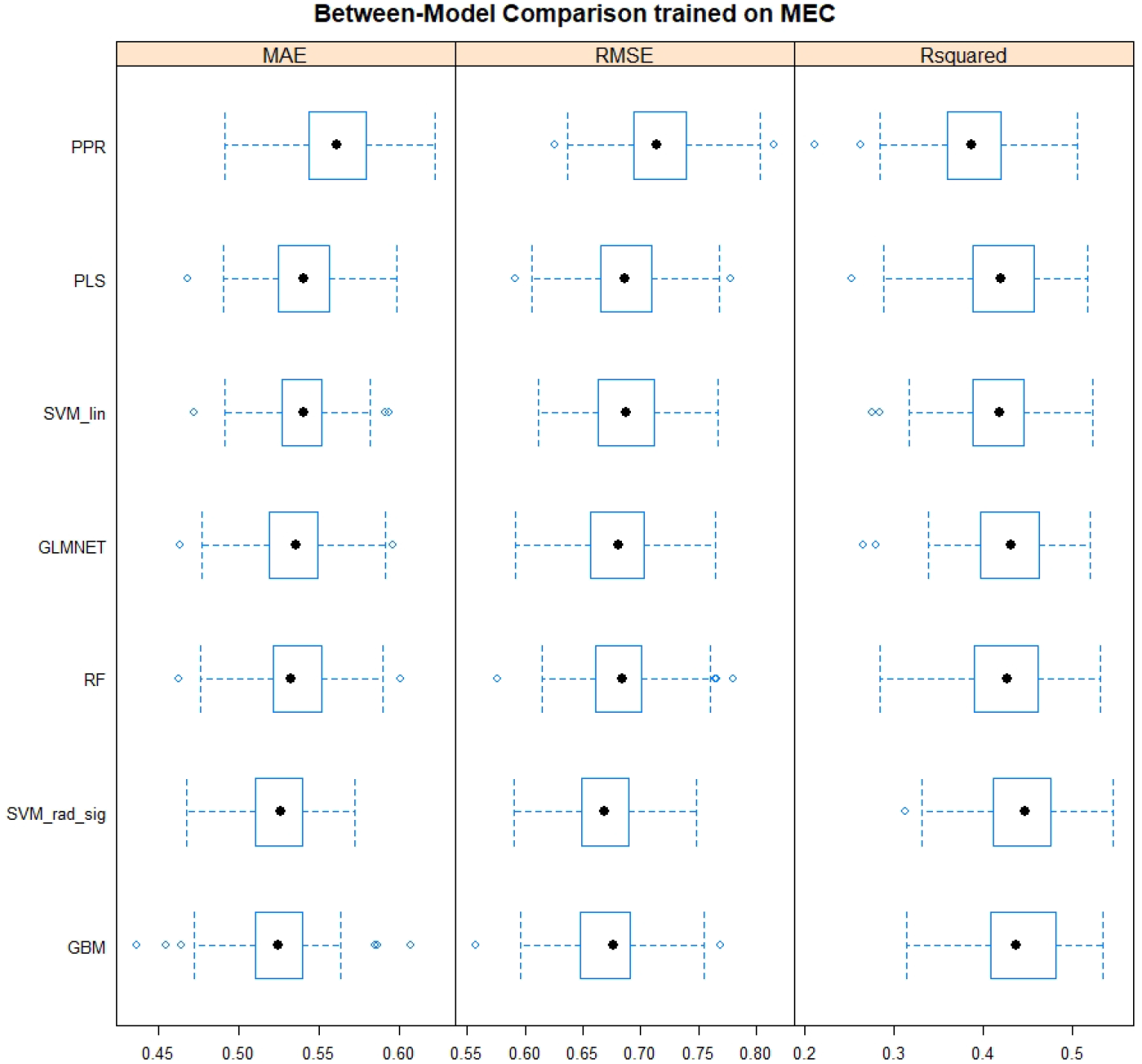
Assessment of the seven models in the training data (MEC). Models were trained using project pursuit (PPR), partial least squares (PLS), support vector machine with a linear kernel (SVM lin), elastic net (GLMNET), random forests (RF), support vector machine with a radial basis function kernel (SVM rad sig), and gradient boosting machine (GBM). Model performances were assessed using mean absolute error (MAE), root mean squared error (RMSE), and R Squared. The boxplots summarizes these metrics across 100 cross validation datasets. Performances were similar across the models justifying use of an average of predictions in the ensemble model.

Once the process of training the models was complete, the ensemble model was constructed; for further discussion on ensemble based techniques see [37]. Let 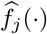 denote the *j*th fitted sub-model. Based on these fitted sub-models, our ensemble is given by 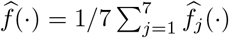. Thus, the trained model can provide predictions of the NMR (*Ŷ*) for a new feature set (**F**) as 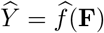. That is, this ensemble provides predictions by averaging the predictions of the individual sub-models. Proceeding in this fashion we obtain more reliable predictive performance than could be obtained from any one of the component models alone. To examine the out of sample performance of our trained ensemble, we use it to predict the NMR for the subjects in the CENIC, HSS, and METS studies. Supplementary Figure S3 provides the predicted vs. the actual NMR across all four studies for each of the seven models and Table 5.3 provides the correlations between predicted and actual NMR by study and model stratified by estimated genomic ancestry. Figure 2 aggregates the predictions from each model to form an ensemble based prediction.

**Figure 2.**
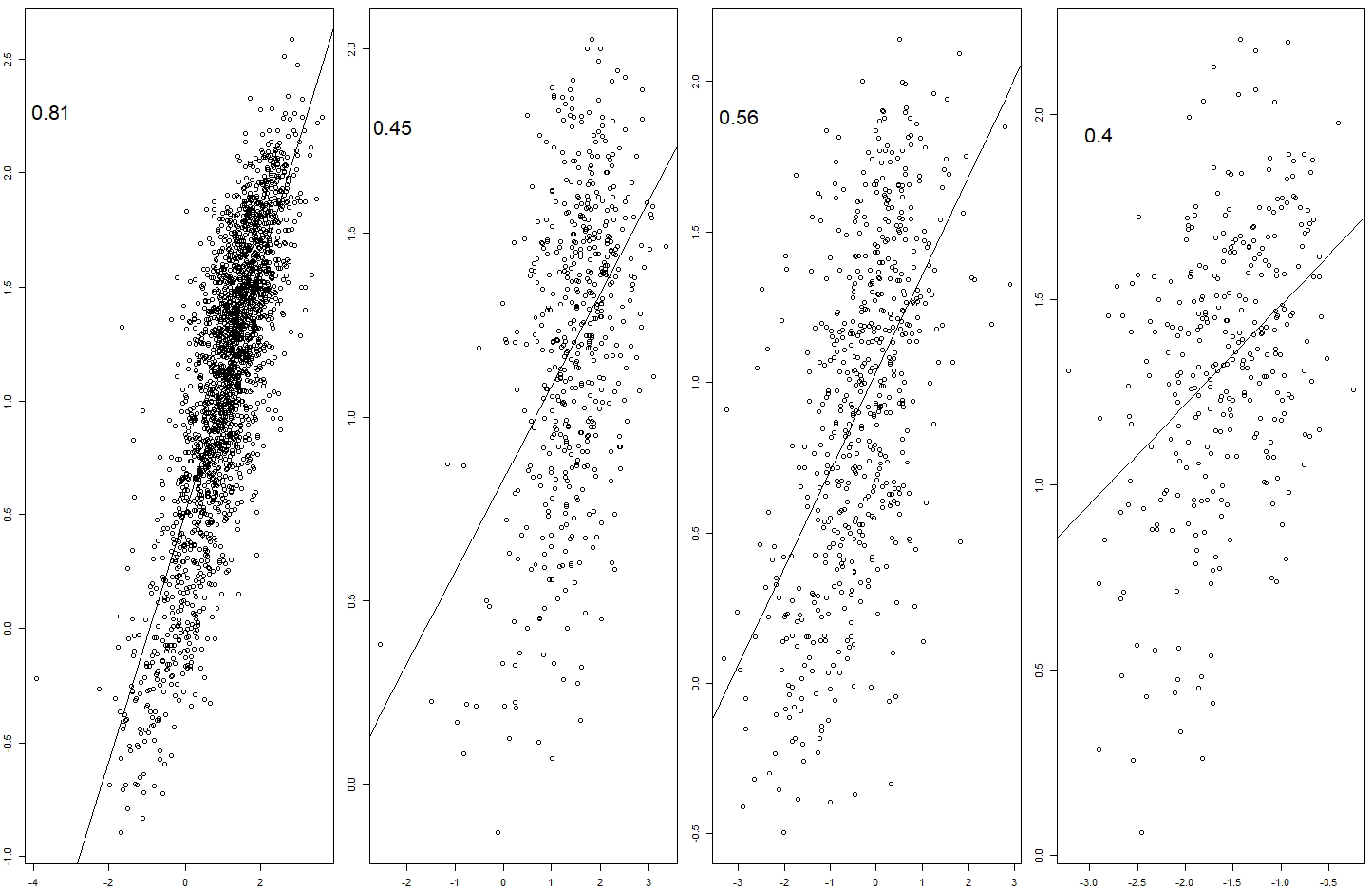
Observed versus predicted NMR values for the training (MEC) and validation (CENIC, HSS, and METS) data. The predicted NMR is the averages of the predictions from the seven models (i.e., the ensemble model). The correlation between these values are displayed to the upper left of each scatterplot. The distribution of NMRs were different across studies, yet the correlations were still strong in the validation datasets.

## 3 Results

### 3.1 Participant Characteristics

The characteristics of the participants for each study are presented in Table 5.3. The MEC and HSS individuals were older on average than CENIC and the METS (mean ages of 64, 61, 43, and 34 respectively). Participants of CENIC had a greater proportion of males than the other studies (59% versus 49% (HSS), 45% (METS), and 46% (MEC)) and larger body mass. The METS study had a mix of smokers (45%) and nonsmokers, whereas the other study participants were all smokers.

The distribution of self-reported races varied by study and as expected corresponded to estimated ancestry proportions (see Supplementary Table 4.2). The MEC included African American (16%), Japanese American (30%), Native Hawaiian/Pacific Islander (14%), Latino (20%), and White (20%) smokers. The METS were comprised of self-reported African American (16%), Asian American (16%), and White (68%) participants. The HSS had nearly equal proportions of Japanese American, Native Hawaiian/Pacific Islander, and White smokers. The smokers in CENIC were mostly White (72%) and African American (21%).

The distributions of natural log NMRs varied by study (Supplementary Figure S1) and represented differences in collection source and timing, metabolite measurements, and study patient characteristics. MEC reported the urinary total 3HC to free COT in smokers; CENIC reported urinary total 3HC to free COT in smokers; HSS reported urinary total 3HC to total COT in smokers; and METS reported plasma or salivary 3HC to COT at 6 hours after a fixed dose of nicotine was administered. This precluded stacking the data for model training.

### 3.2 Model development

The distribution of marginal p-values found in the four genome-wide association scans of NMR are provided in Supplementary Figure S2. There does not appear to be any inflation or deflation of the p-values overall (i.e. *λ*’s are close to one). In each study, there were many genetic variants with small p-values; as seen in the the tail of the distributions presented in Supplementary Figure S2. The smallest 200 p-values with allele frequencies > 1% were merged across the studies. After filtering (see Section 2.6.1), there were 263 genetic variants selected as candidates predictors of NMR. The list of markers and their corresponding chromosome, position, and alleles can be found in the Supplementary Files. MEC had the largest and most diverse sample of the studies considered. Given this, we trained the models on the MEC dataset and validated them in the other three studies.

### 3.3 Model performances

Within MEC, model performances were summarized across 100 model fits at their optimal tuning parameter configuration which are presented in Figure 1. These included the mean absolute error (MAE), root mean squared error (RMSE), and the R-squared. Models with lower values of MAE and RMSE achieve higher model accuracy. The R-squared is the proportion of variance explained by the model. As shown in Figure 1, the trained models can explain about half of the variability in NMR. The performances across the models are remarkably similar, with no clear winner or loser. Given this observation, we give each model equal weights in the ensemble model. That is, the ensemble model is simply the average of the predicted NMR values from the seven component models.

The model performances within sample (MEC) and out-of-sample (HSS, METS, CENIC) for each model and the overall ensemble are presented in Supplementary Figure S3 and Figure 2 respectively. Within MEC, the correlations between the observed and predicted NMR ranges from 0.69 to 0.97 depending on the model. The correlation of the ensemble and observed values is 0.81 indicating that averaging the predictions from the member models yields good prediction performance. The models also generalizes well out-of-sample. The correlation between the observed NMR and the predicted NMR from the ensemble are 0.45, 0.56, and 0.4 for CENIC, HSS, and METS respectively (Figure 2).

The correlations for each component model stratified by ancestry are presented in Table 5.3. Overall, the ensemble model performed well across ancestries in the MEC (0.76, 0.82, and 0.67 for African, Asian, and European ancestries respectively). In the validation studies, the best ensemble correlations for African ancestry was 0.52 in the METS, for Asian ancestry was 0.61 in CENIC, and for European ancestry was 0.46 in CENIC.

### 3.4 Variable Importance

Variable importance is an indicator of how much a candidate predictor contributes to a model on a scale of zero to 100. These are presented for the seven models trained in the MEC data in Supplementary Figure S4. In examining the patterns, there was consensus among the models on the importance of many of the variables in predicting NMR yet diversity in the variables each models deemed important. Given this observation and the performances of the models being rather similar, the panel of 263 genetic variants seems adequate. We next examine the favorites by ranking variables by importance for each model and counting how many times each variable occurs in the top 20 (Figure 3). Asian ancestry and rs56113850 were highly relevant to the prediction of NMR in all models. rs11878604 was also important in predicting NMR (six models). Additionally, African ancestry, age, and 24 genetic variants were of top importance in more than two models.

**Figure 3.**
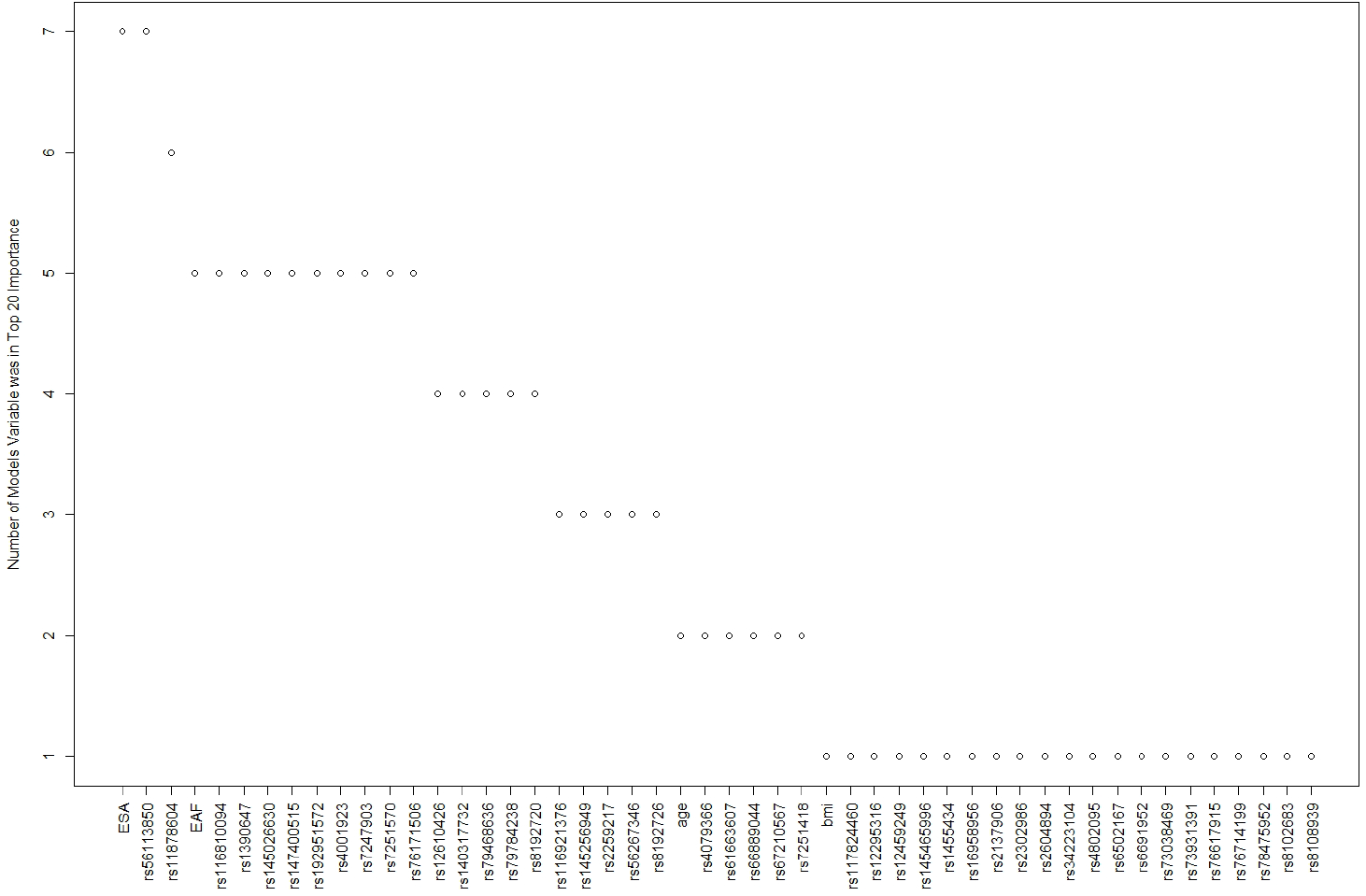
Summary of the most important candidate variables in the NMR models. Variable importance was ranked for each of the seven models trained on the MEC data. The number of times each variable occurred in the top 20 for each model was enumerated. Asian ancestry and rs56113850 was an influential variable in all the models.

## 4 Discussion

The NMR is an important biomarker for selecting the optimal intervention for smokers seeking to quit and for evaluating the risks of tobacco use. We selected a panel of 263 genetic markers from genome-wide analyses in four multiethnic dataset, trained the models in the largest, most ancestrally diverse dataset, and validated models in three additional multiethnic datasets. In summary, we have created an ensemble model for estimating the NMR across ancestries from easily obtainable genotypes and covariates (i.e., age, height, weight, gender, and race/ancestry). This work also provides a methodological framework for developing other genomic-based assessments of heritable biomarkers.

Diversity was a strengths in our approach and findings. The American individuals used in model training and validation studies represented diverse ancestries, composed of Asian, African, and European backgrounds. This enabled us to detect genetic variants related to nicotine metabolism that we would not have selected otherwise and train models that generalize to new observations. This is highlighted in Table 5.3 by the out-of-sample performances of the models in CENIC, HSS, and METS across ancestries. We also adopted an ensemble based approach that averages NMR predictions from seven different component models. While performances of these seven models were similar, there was variation in how the candidate variables contribute to the model (Supplementary Figure S4). This diversity in solutions is a strength, enabling more reliable predictions in the ensemble model.

The most important predictors in these models combine many of the findings in the literature on the genetics of the NMR. Of the 26 genetic variants flagged in more than one model (Figure 3), 7 had entries in the NHGRI-EBI Catalog of human genome-wide association studies [38]. The marker rs56113850 was associated with nicotine metabolism at genome-wide significance in smokers of European ancestry [10, 11]. In smokers of African ancestry, the marker rs11878604 was associated with NMR at genome-wide significance and identified as an independent signal [39]. This marker was also top ranked in African American female smokers [40]. The marker rs12459249 was associated at genome-wide significance and top ranked in the METS meta-GWAS [26], and was an independent signal identified in conditional analysis of African American treatment seeking smokers [39]. Markers rs11878604 and rs116921376 have been implicated in tobacco-related consequences (lung cancer and chronic obstructive pulmonary disease) [41]. The marker rs56267346 has been identified as playing a role in caffeine metabolism, which involves *CYP2A6* [42]. rs8192726 has been shown to be related to cigarettes per day in a genomic study of 1.2 million individuals [43]. These variants are all located at or near the *CYP2A6* gene on chromosome 19. Age and BMI have previously been shown to influence the NMR in treatment-seeking smokers [44]. Genomic estimated Asian and African ancestries were important in five and seven models, respectively, and have been shown to influence nicotine metabolism [45].

Differences in NMR measurements prevented us from stacking the data across studies. These differences included the type of biospecimens collected (urine, blood, saliva), the timing of collection, and the metabolite measurement procedures. For example, in the METS, participants were administered fixed doses of labeled nicotine and cotinine and biospecimens were collected at regular intervals, with the 6 hour collection being used to assess NMR. However, this timing does not allow the metabolite ratio to reach a steady state, thus the NMR is under reported. Additionally, the HSS measured total cotinine, which is more sensitive to variations in cotinine glucuronidation than free cotinine [8]. These differences however provided a unique opportunity for validation since while different, we know these NMRs are correlated. As such, in our analysis, we trained on the largest and most diverse data (MEC), and were able to validate the models in the other three studies (METS, CENIC, and HSS). While the correlation between the observed and predicted NMRs were the strongest in the training set as expected (0.81 for the MEC ensemble model), the models also predicted NMRs out of sample (correlations of 0.45, 0.58, and 0.40 for CENIC, HSS, and METS respectively).

As noted, we trained the models on the largest multiethnic sample. However, MEC was genotyped using an older genotype array (Illumina Human1M-Duo BeadChip), while the other studies were genotyped with the Smokescreen Genotyping Array, designed with more markers within genes regions related to nicotine metabolism and smoking-related behaviors outcomes [26, 14]. While all studies were imputed to the 1000 Genomes Project, there were more poor quality genotypes (typed or imputations) in the older studies MEC (23 markers) and METS (28 markers) than the newly genotyped studies (5 markers in CENIC and 12 markers in HSS). Additionally there were *CYP2A6* variants not considered in this analysis. For example, there are functional structural variations in *CYP2A6* not directly typed on genomic arrays (i.e., copy number variants, gene duplication, deletions, and translocations). There were also environmental factors that influence NMR that were not considered here, e.g., estrogen, comorbidites and diet [1]. These factors (and interactions with such factors) can and should be incorporated into future studies to improve prediction.

### 4.1 Validation with smoking dosage in the HSS and CENIC

The rate of nicotine metabolism influences how much nicotine an individual is exposed to (i.e., nicotine dosage) and consequently risk of lung cancer [3]. Nicotine equivalents is the sum of nicotine and nicotine metabolites, and offers a more precise measure of nicotine intake than self-reported cigarettes per day. To link predicted nicotine metabolism to nicotine exposure, we took the NMR predictions for HSS and CENIC using the MEC trained ensemble model, and examined their relationship to nicotine equivalents. We found that the predicted NMRs were strongly associated with nicotine equivalents in both studies (*p* = 0.000327 and *p* = 1.6*E* − 7 in CENIC and HSS respectively). This indicates that predicting how an individual metabolizes nicotine could be used to quantify their nicotine exposure and tobacco-attributable disease risk.

### 4.2 Implications and Future Work

Direct measurement of the NMR has its challenges. For example, individuals must be actively using nicotine-containing products and there are issues related to measurement and sample collection. We offer an approach where easy to obtain genotypes and basic demographics could be used to characterize how a current or previous tobacco user metabolizes nicotine. Genotypes could be obtained by inexpensive genotyping platforms and paired with popular saliva DNA collection kits. The knowledge of how an individual metabolizes nicotine could be used to help select the optimal path to reducing or quitting tobacco use, as well as, evaluating risks of tobacco-related diseases and comorbidities.

While additional work is needed to optimize the predictive models using larger population representative samples with genotypes and both plasma and urine nicotine metabolites, the presented models may be immediately used to predict nicotine metabolism in newly collected or existing DNA samples, or from existing genomic data. These predictions in turn can be linked to clinical outcomes. For example probabilistic models could be built that relate the predicted NMR to the likelihood of smoking cessation or response to different treatment options. This could lead to the identification of NMR cut-points that could be used to guide subject specific treatment paths.

## Data Availability

The datasets generated during and/or analysed during the current study are available from the corresponding author on reasonable request.

## 5 Declarations

### 5.1 Ethics approval and consent to participate

Written informed consent was obtained from all participants. The research described herein received approvals from the Institutional Review Boards of BioRealm, Oregon Research Institute, and the University of Hawaii.

### 5.2 Consent for publication

Not applicable.

## Competing interests

JWB and CME are members and employees of BioRealm LLC. AWB is an employee of Oregon Research Institute and ORI Community and Evaluation Services, and serves as a Scientific Advisor and Consultant to BioRealm LLC. JWB, CSM and AWB are co-inventors on a related patent application “Biosignature Discovery for Substance Use Disorder Using Statistical Learning”, assigned to BioRealm LLC. BioRealm LLC offers genotyping and data analysis services. Other authors declare that they have no competing interests.

## Funding

This study was funded by the National Institute on Alcohol Abuse and Alcoholism (R44 AA027675) and the National Institute on Drug Abuse (R43 DA041211). The sponsors had no role in the analysis of data, writing of the report, or in the decision to submit the paper for publication.

## Author’s contributions

All authors contributed to preparing this manuscript. AWB, SLP, SEM prepared data and samples; JWB, AWB, and CME performed data management; and JWB and CME performed data analysis.

## Acknowledgements

The authors thank participants, staff and Investigators of the MEC, the CENIC, the HSS and the METS. The METS were supported by: the National Institute on Drug Abuse Pharmacokinetics and Pharmacodynamics of Nicotine (DA002277, PI: Neal L Benowitz), Young Adult Substance Use-Predictors and Consequences (DA003706, PI: Hy Hops), Pharmacokinetics of Nicotine in Twins (DA011170, PI: Gary E Swan), Pharmacogenetics of Nicotine Addiction Treatment Consortium (DA020830, PI: Neal L Benowitz; MPI: Rachel F Tyndale, and Caryn Lerman); and, by the Tobacco-Related Disease Research Program of the University of California: Nicotine Metabolism in Families (7PT-2004, PI: Neal L Benowitz). The MEC was supported by the National Cancer Institute (U01 CA164973 and P01 CA138338). CENIC was supported by a grant from the National Institute on Drug Abuse and the Food and Drug Administration Center for Tobacco Products (U54 DA031659). The HSS was supported by National Cancer Institute (R01 CA 85997). We would like to acknowledge BioRealm LLC team (https://biorealm.ai) for supporting project workflows and computation and IBX (http://ibx.bio) for sample processing.

## Tables

**Table 1.** Participant characteristics for the training data (MEC) and the three validation datasets (CENIC, HSS, and METS)

|  | MEC<br>(N=2239) | CENIC<br>(N=515) | HSS<br>(N=580) | METS<br>(N=315) |
| --- | --- | --- | --- | --- |
| **Age** |  |  |  |  |
| Mean (SD) | 63.9 (7.19) | 43.4 (13.2) | 60.6 (9.40) | 33.8 (10.9) |
| Median [Min, Max] | 63.0 [45.0, 86.0] | 44.0 [18.0, 75.0] | 60.6 [19.6, 83.2] | 30.0 [18.0, 69.0] |
| **Body Mass Index** |  |  |  |  |
| Mean (SD) | 26.3 (5.31) | 30.0 (6.70) | 27.1 (6.10) | 25.6 (4.75) |
| Median [Min, Max] | 25.6 [11.3, 62.8] | 29.2 [15.2, 56.0] | 26.1 [14.4, 54.8] | 24.9 [15.9, 49.1] |
| Missing | 0 (0%) | 3.00 (0.5%) | 0 (0%) | 0 (0%) |
| **Gender** |  |  |  |  |
| Male | 1040 (46.4%) | 306 (59.4%) | 284 (49.0%) | 141 (44.8%) |
| Female | 1199 (53.6%) | 209 (40.6%) | 296 (51.0%) | 174 (55.2%) |
| **Current Smoker** |  |  |  |  |
| Yes | 2239 (100%) | 515 (100%) | 580 (100%) | 120 (38.1%) |
| No | 0 (0%) | 0 (0%) | 0 (0%) | 195 (61.9%) |
| **Self Reported Race** |  |  |  |  |
| African American | 364 (16.3%) | 107 (20.8%) | 0 (0%) | 49 (15.6%) |
| American Indian/Alaskan Native | 0 (0%) | 5 (1.0%) | 0 (0%) | 0 (0%) |
| Asian American | 0 (0%) | 6 (1.2%) | 0 (0%) | 51 (16.2%) |
| Multirace | 0 (0%) | 25 (4.9%) | 0 (0%) | 0 (0%) |
| White | 437 (19.5%) | 372 (72.2%) | 197 (34.0%) | 215 (68.3%) |
| Japanese American | 674 (30.1%) | 0 (0%) | 191 (32.9%) | 0 (0%) |
| Native Hawaiian/Pacific Islander | 311 (13.9%) | 0 (0%) | 192 (33.1%) | 0 (0%) |
| Latino | 453 (20.2%) | 0 (0%) | 0 (0%) | 0 (0%) |
| **Natural log Nicotine Metabolite Ratio** |  |  |  |  |
| Mean (SD) | 1.11 (0.898) | 1.50 (0.720) | -0.324 (0.959) | -1.58 (0.548) |
| Median [Min, Max] | 1.20 [-3.91, 3.60] | 1.54 [-2.54, 3.35] | -0.267 [-3.30, 2.90] | -1.57 [-3.22, -0.240] |

**Table 2.**
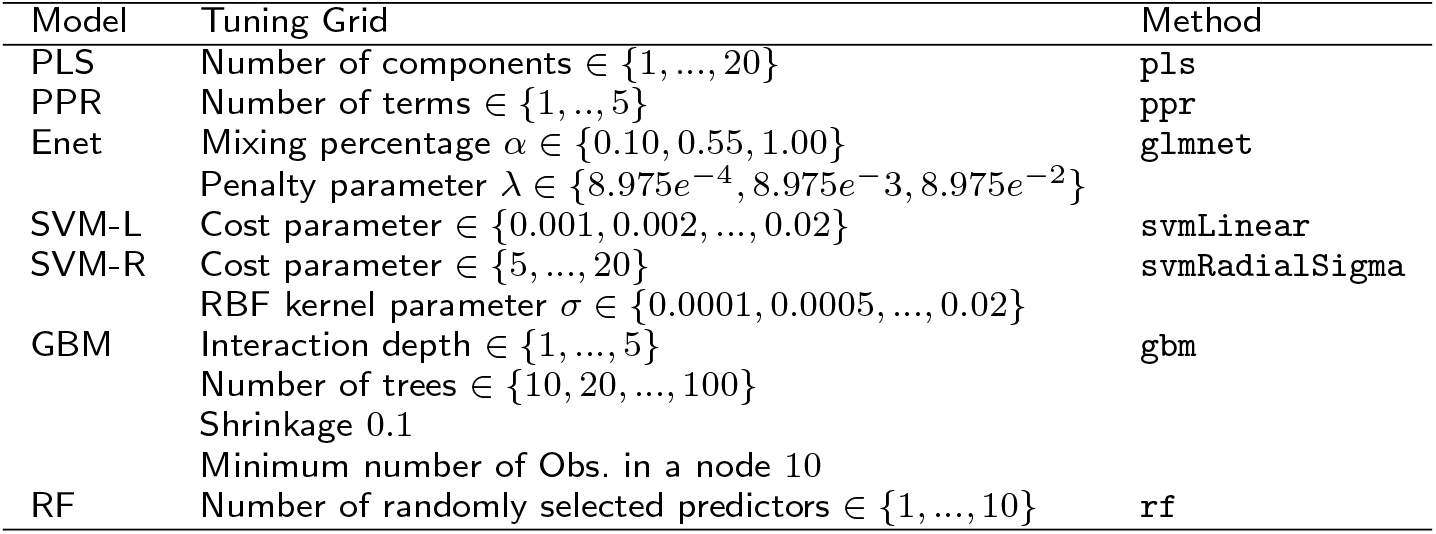
Fitting method and tuning parameter configurations. Provided are the considered training parameters for partial least squares (PLS), project pursuit (PPR), elastic net (ENet), support vector machine with a linear kernel (SVM-L), support vector machine with a radial basis function kernel (SVM-R), gradient boosting machine (GBM), and random forests (RF). Also provided are the model fitting methods.

| Model | Tuning Grid | Method |
| --- | --- | --- |
| PLS | Number of components $\in \{1, \dots, 20\}$ | pls |
| PPR | Number of terms $\in \{1, \dots, 5\}$ | ppr |
| Enet | Mixing percentage $\alpha \in \{0.10, 0.55, 1.00\}$<br>Penalty parameter $\lambda \in \{8.975e^{-4}, 8.975e^{-3}, 8.975e^{-2}\}$ | glmnet |
| SVM-L | Cost parameter $\in \{0.001, 0.002, \dots, 0.02\}$ | svmLinear |
| SVM-R | Cost parameter $\in \{5, \dots, 20\}$<br>RBF kernel parameter $\sigma \in \{0.0001, 0.0005, \dots, 0.02\}$ | svmRadialSigma |
| GBM | Interaction depth $\in \{1, \dots, 5\}$<br>Number of trees $\in \{10, 20, \dots, 100\}$<br>Shrinkage 0.1<br>Minimum number of Obs. in a node 10 | gbm |
| RF | Number of randomly selected predictors $\in \{1, \dots, 10\}$ | rf |

**Table 3.** Correlations between predicted and observed NMRs by study, ancestry, and model. The correlations between predicted and observed NMRs were summarized overall and by genomic ancestry (ancestry proportion > 0.5). The MEC was the largest and most diverse sample and was used for training using partial least squares (PLS), project pursuit (PPR), elastic net (ENet), support vector machine with a linear kernel (SVM-L), support vector machine with a radial basis function kernel (SVM-R), gradient boosting machine (GBM), and random forests (RF). Predictions from these seven models were averaged in an ensemble model. The MEC trained models were applied to CENIC, HSS, and METs for validation.

| **MEC (Training)** | PLS | PPR | ENet | SVM-L | SVM-R | GBM | RF | Ensemble | N |
| --- | --- | --- | --- | --- | --- | --- | --- | --- | --- |
| African | 0.60 | 0.67 | 0.58 | 0.75 | 0.58 | 0.76 | 0.97 | 0.76 | 342 |
| Asian | 0.71 | 0.76 | 0.70 | 0.79 | 0.69 | 0.81 | 0.97 | 0.82 | 995 |
| European | 0.50 | 0.56 | 0.49 | 0.63 | 0.48 | 0.64 | 0.97 | 0.67 | 902 |
| Overall | 0.71 | 0.75 | 0.70 | 0.78 | 0.69 | 0.79 | 0.97 | 0.81 | 2239 |
| **CENIC (Validation)** |  |  |  |  |  |  |  |  |  |
| African | 0.35 | 0.33 | 0.32 | 0.33 | 0.32 | 0.41 | 0.41 | 0.37 | 111 |
| Asian | 0.71 | 0.28 | 0.68 | 0.60 | 0.67 | 0.51 | 0.50 | 0.61 | 9 |
| European | 0.42 | 0.38 | 0.43 | 0.41 | 0.37 | 0.52 | 0.47 | 0.46 | 395 |
| Overall | 0.42 | 0.37 | 0.41 | 0.39 | 0.37 | 0.50 | 0.45 | 0.45 | 515 |
| **HSS (Validation)** |  |  |  |  |  |  |  |  |  |
| African |  |  |  |  |  |  |  |  | 1 |
| Asian | 0.51 | 0.29 | 0.55 | 0.59 | 0.53 | 0.55 | 0.58 | 0.56 | 308 |
| European | 0.42 | 0.33 | 0.42 | 0.37 | 0.36 | 0.42 | 0.39 | 0.43 | 271 |
| Overall | 0.53 | 0.37 | 0.56 | 0.56 | 0.53 | 0.55 | 0.56 | 0.56 | 580 |
| **METS (Validation)** |  |  |  |  |  |  |  |  |  |
| African | 0.43 | 0.30 | 0.50 | 0.55 | 0.41 | 0.46 | 0.45 | 0.52 | 48 |
| Asian | 0.37 | 0.03 | 0.39 | 0.47 | 0.41 | 0.47 | 0.47 | 0.43 | 51 |
| European | 0.38 | 0.10 | 0.40 | 0.45 | 0.42 | 0.44 | 0.41 | 0.43 | 216 |
| Overall | 0.36 | 0.09 | 0.42 | 0.42 | 0.39 | 0.45 | 0.45 | 0.40 | 315 |

## Additional Files

Additional file 1 — Supplementary Figures

Additional file 2 — Supplementary Table

Additional file 3 — List of genetic variants nominated for prediction of nicotine metabolism

This file contains the rs number, chromosome, hg19 position, reference allele, and alternative allele for the 263 genetic variants selected as candidates for prediction of the nicotine metabolite ratio.

## Supplementary Figures

**Figure S1.**
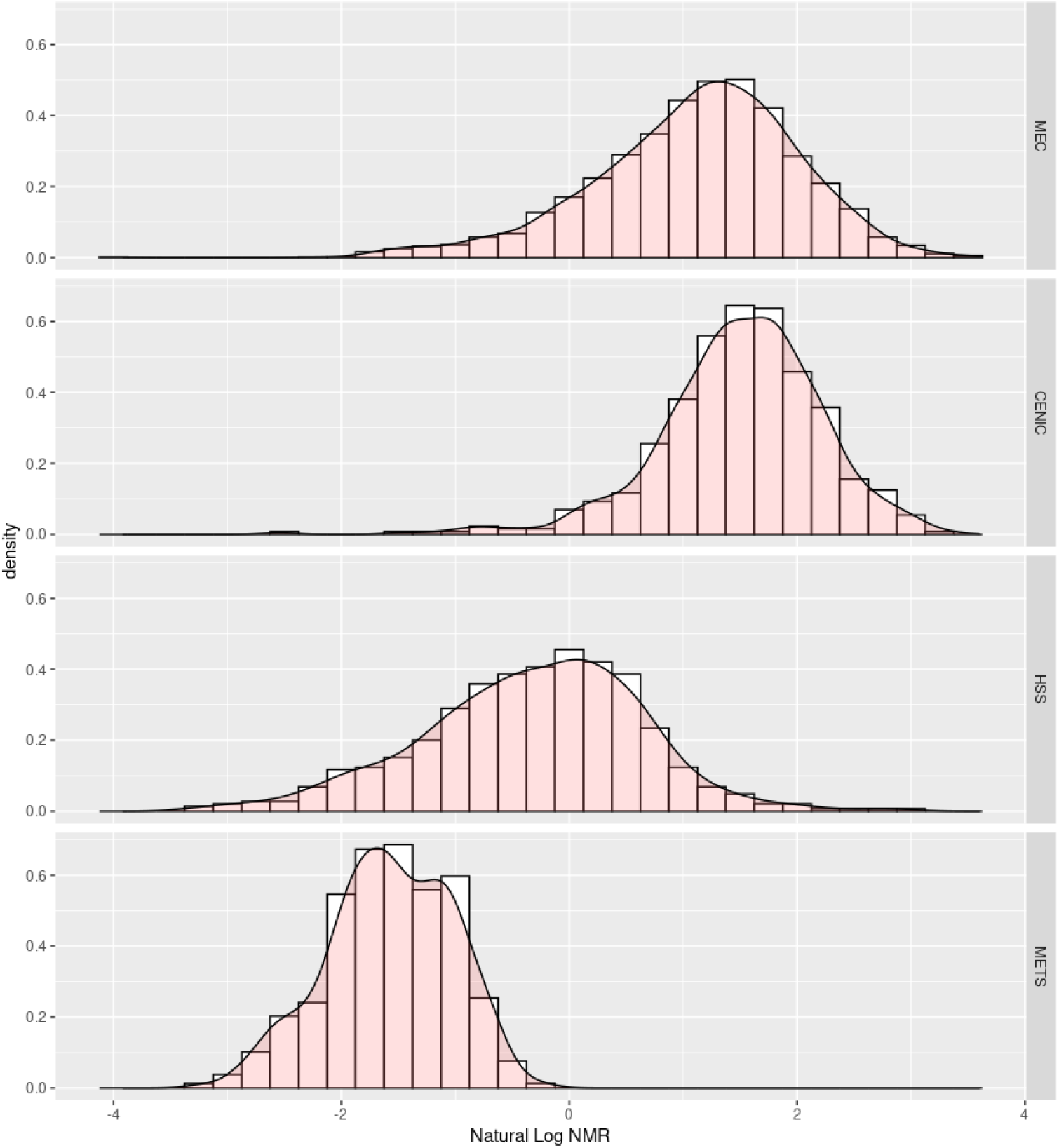
Distribution of the natural log nicotine metabolite ratio by study. The MEC and CENIC NMR measures were total *trans*-3’-hydroxycotinine (3HC) to free cotinine (COT) in urine. The HSS NMR measure was total 3HC to total COT in urine. The METS NMR measure was 3HC to free COT in plasma or saliva six hours after administration of a fixed dose of deuterium-labeled nicotine and cotinine.

**Figure S2.**
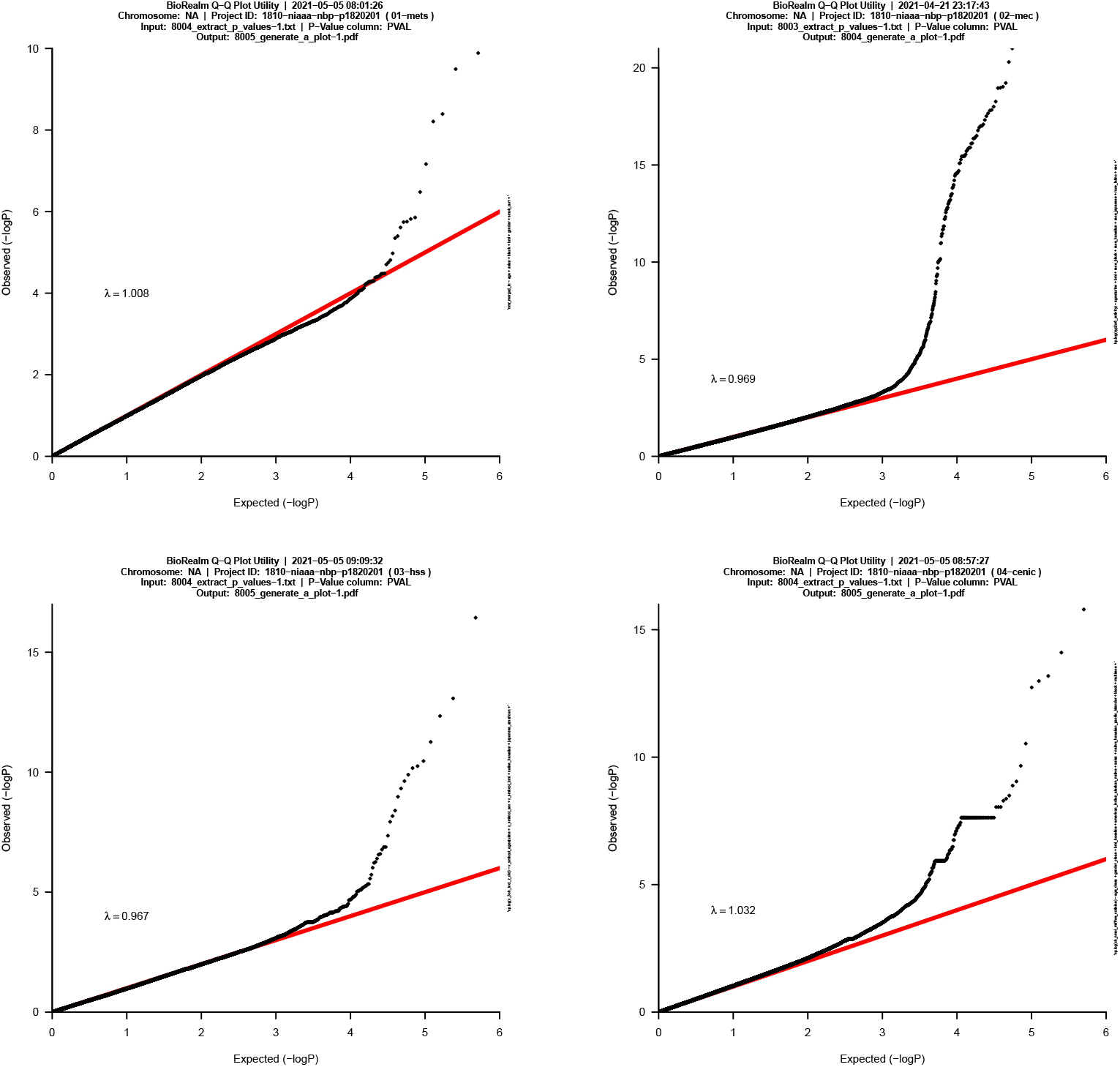
Observed versus expected distribution of marginal p-values from four genome wide association scans of NMR. Studies from left to right: METS, MEC, HSS, and CENIC. Each model is adjusted for age, sex, BMI, race, and 50 principal components. There is no evidence of systematic inflation or deflation of the p-values. The tails of the distributions represent genetic variants nominated as candidate variables for NMR prediction models.

**Figure S3.**
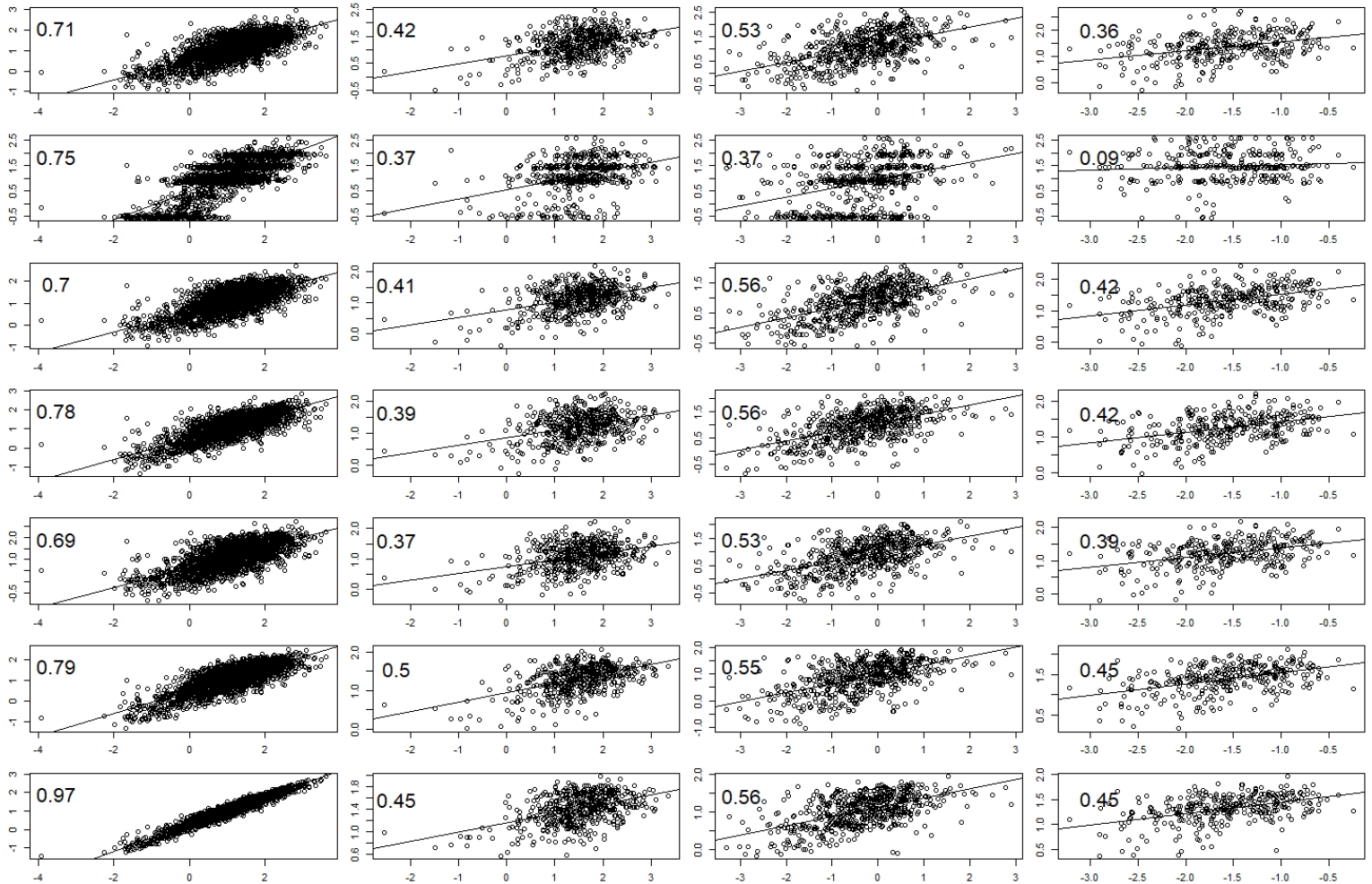
Plots of predicted versus observed NMR values by four studies (columns) and seven machine learning models (rows). The first column present scatterplots for the training dataset (MEC). The next three columns are the results for the validation datasets (CENIC, HSS, and METS). The models from top to bottom are partial least squares (PLS), project pursuit (PPR), elastic net (ENet), support vector machine with a linear kernel (SVM-L), support vector machine with a radial basis function kernel (SVM-R), gradient boosting machine (GBM), and random forests (RF). The seven models have similar performances in the training data, and thus have equal weights in the ensemble model. It is important to note that the distributions of NMR differ across the studies due to differences in how NMR was collected, measured, and patient characteristics. Yet, correlations are still strong in applying the models trained in MEC to the other datasets.

**Figure S4.**
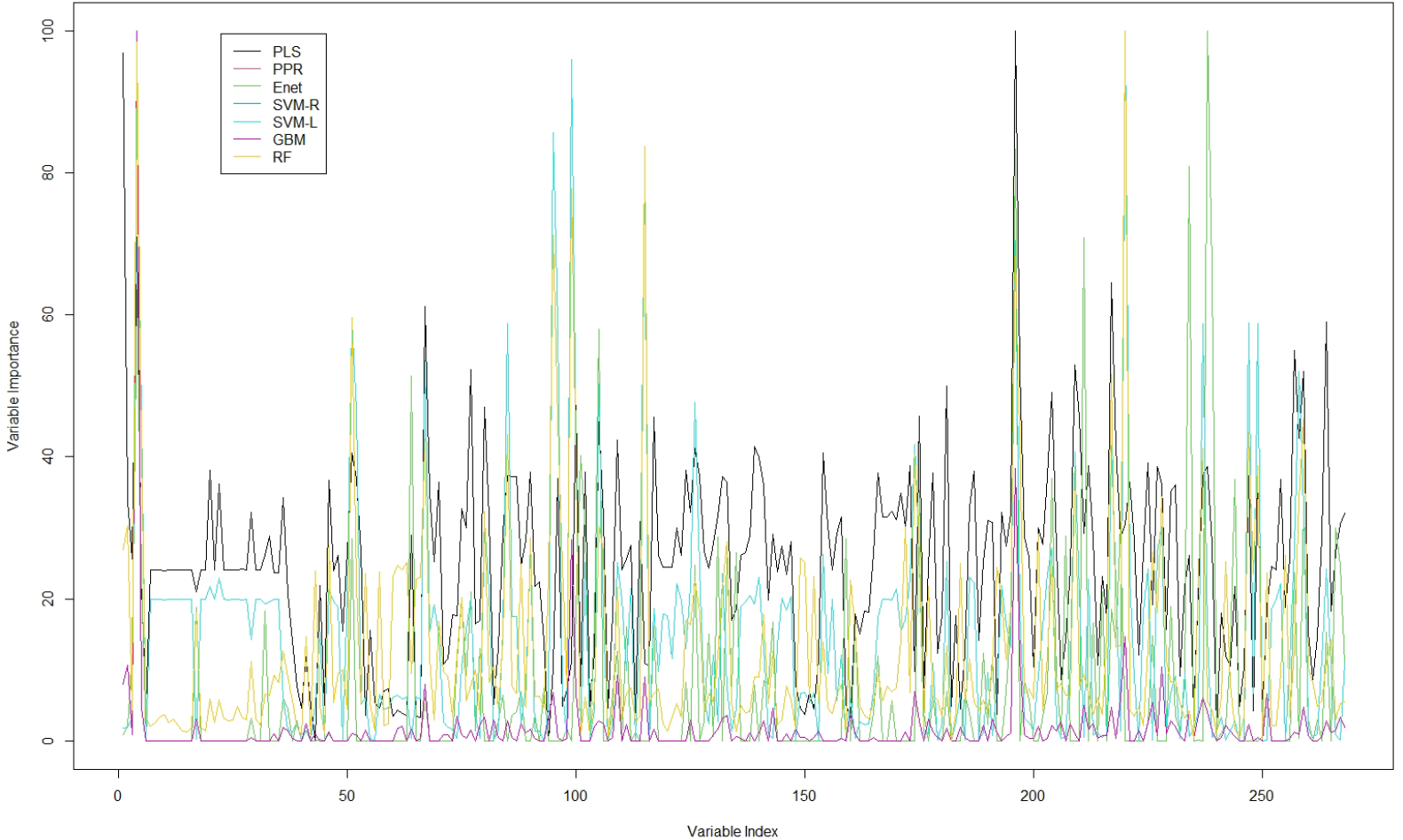
Candidate variable importance for seven machine learning models trained on the MEC dataset. The 263 genomic variables and covariates considered by each modeling approach are indexed along the x-axis. The influence each variable has on NMR is provided on a scale of 0 to 100 for each model, with higher variable importance indicating greater influence on predicted NMR. There was concordance among the models on the importance of many variables, yet diversity among the models. This property is desirable for ensemble predictions as each component model is using different features of the data to yield similar overall prediction performances.

## Supplementary Tables

**Table S1.**
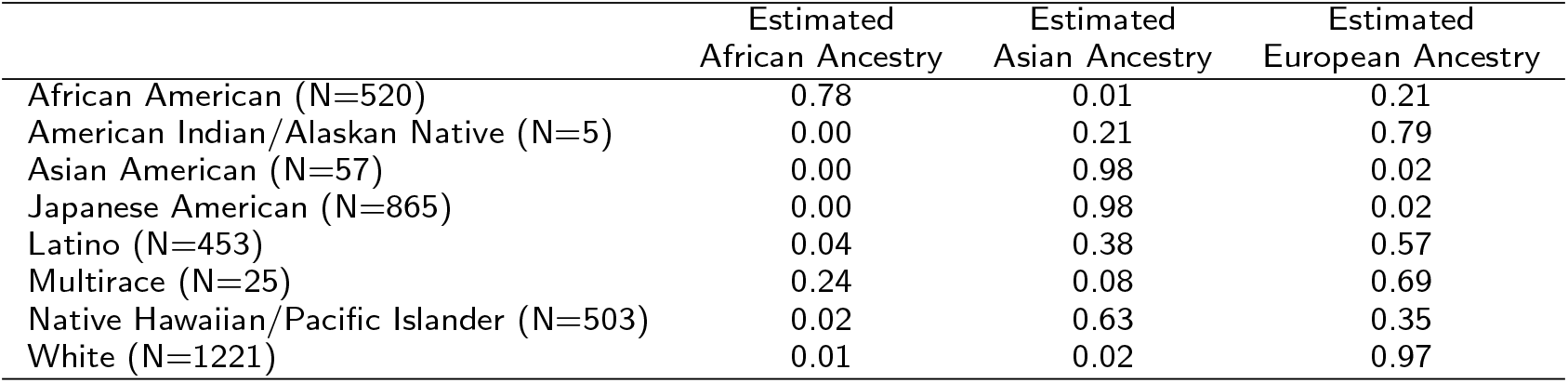
Mean genomic ancestry proportions by self-reported race. Genomic ancestries were estimated by applying fastSTRUCTURE to genotypes for 5516 ancestry informative markers extracted from each study and 1000 Genomes Project samples with known population labels. Ancestry proportions were averaged for each self-reported race.

|  | Estimated<br>African Ancestry | Estimated<br>Asian Ancestry | Estimated<br>European Ancestry |
| --- | --- | --- | --- |
| African American (N=520) | 0.78 | 0.01 | 0.21 |
| American Indian/Alaskan Native (N=5) | 0.00 | 0.21 | 0.79 |
| Asian American (N=57) | 0.00 | 0.98 | 0.02 |
| Japanese American (N=865) | 0.00 | 0.98 | 0.02 |
| Latino (N=453) | 0.04 | 0.38 | 0.57 |
| Multirace (N=25) | 0.24 | 0.08 | 0.69 |
| Native Hawaiian/Pacific Islander (N=503) | 0.02 | 0.63 | 0.35 |
| White (N=1221) | 0.01 | 0.02 | 0.97 |

